# Comparing AI- versus clinician-authored summaries of simulated primary care electronic health records

**DOI:** 10.1101/2025.02.21.25322674

**Authors:** Lara Shemtob, Abdullah Nouri, Adam Sullivan, Connor S Qiu, Jonathan Martin, Martha Martin, Sara Noden, Tanveer Rob, Ana Luisa Neves, Azeem Majeed, Jonathan Clarke, Thomas Beaney

## Abstract

**Objective:** To compare clinical summaries generated from simulated patient primary care electronic health records (EHRs) by ChatGPT-4 to summaries generated by clinicians on multiple domains of quality including utility, concision, accuracy and bias.

**Materials and Methods:** Seven primary care physicians generated 70 simulated patient EHR notes, each representing 10 appointments with the practice over at least two years. Each record was summarised by a different clinician and by ChatGPT-4. AI- and clinician-authored summaries were rated blind by clinicians according to eight domains of quality and an overall rating.

**Results:** The median time taken for a clinician to read through and assimilate the information in the EHRs before summarising, was seven minutes. Clinicians rated clinician-authored summaries higher than AI-authored summaries overall (7.39 versus 7.00 out of 10; p=0.02), but with greater variability in clinician-authored summary ratings. AI and clinician-authored summaries had similar accuracy and AI-authored summaries were less likely to omit important information and more likely to use patient-friendly language.

**Discussion:** Although AI-authored summaries were rated slightly lower overall compared with clinician-authored summaries, they demonstrated similar accuracy and greater consistency. This demonstrates potential applications for generating summaries in primary care, particularly in the context of the substantial time taken for clinicians to undertake this work.

**Conclusion:** The results suggest the feasibility, utility and acceptability of using AI-authored summaries to integrate into EHRs to support clinicians in primary care. AI summarisation tools have the potential to improve healthcare productivity, including by enabling clinicians to spend more time on direct patient care.

## Background and Significance

Electronic health records (EHRs) contain a huge volume of health-related data. While some data may be entered as ‘structured’ clinical codes, for example using an alphanumeric code to signify the presence of a diagnosis, this reflects only a small proportion of the information recorded in EHR data in the form of ‘unstructured’, or ‘free text’ data, which can amount to thousands of words (1,2). Reading this information in preparation for a consultation is a laborious and time-consuming task for clinical staff (1). Clinicians spend a significant amount of time interacting with EHRs, (3) and sometimes, more time is spent interacting with EHRs on administrative tasks than is spent delivering direct patient care (4). Even when search functions are available to retrieve entries with text relevant to a particular problem, relevant documentation can be missed, leading to safety issues (5). Analyses of patient safety reports have found that difficulties in easily combining and interrogating documentation from patient encounters can contribute to safety events (6).

Recently, Large Language Models (LLMs) have rapidly evolved and are capable of wide-ranging applications using text data, such as question-and-answering, text generation, translation and summarisation. If LLMs could accurately summarise the extensive information recorded in EHRs, this could improve relevant information retrieval, enhance informational continuity and potentially reduce clinical errors (7). Effective summarisation could allow for provision of accessible summaries to patients of their own EHR notes, or generation of summaries of a patient’s medical history when referring to another healthcare organisation, improving continuity between providers. Summaries of a patient’s relevant medical history could also help healthcare professionals to prepare when they review EHR notes before a consultation. To our knowledge, such applications of LLMs are not currently in use but have the potential to reduce clinical workloads while also reducing the risk of vital information being missed.

Existing literature reporting on the application of LLMs within EHRs across healthcare settings largely focuses on the ability of the technology to undertake various information extraction and prediction tasks (8). Much of this work has taken place in hospital settings, (9) rather than primary care, where EHRs are arguably richer, including a longitudinal history documenting medical diagnoses, prescriptions and test results across a person’s life (10)(11). One study that compared using clinician versus AI suggestions from ChatGPT to optimise clinical decision support in primary care EHRs found those from ChatGPT were evaluated as highly understandable and relevant (12). Furthermore, ChatGPT can effectively undertake summarisation of clinical dialogues in primary care, as evaluated by human experts (13). Other work demonstrates that different LLMs may outperform humans across summarisation tasks, including summarising radiology reports and extracting key information from secondary care notes (14,15). To our knowledge, the use of LLMs to summarise free text written data in primary care EHRs has not been investigated. In this study, we compare the quality of EHR summaries relevant to a clinician before consulting with a patient, generated by AI, to those generated by clinicians.

## Materials and Methods

### Overview of study design

This is a comparative study of the quality of clinical summaries authored by ChatGPT-4 versus clinicians, with clinicians blinded to the source of the summary during evaluation. Figure 1 summarises the study design. Seven primary care clinicians each generated ten simulated patient EHR notes representing the 10 most recent longitudinal contacts with the practice over at least the last two years, resulting in 70 records. These simulated EHRs were generated using templates representative of demographics and chronic condition prevalence in the population of patients registered with general practices in England. Each record was summarised independently by a primary care clinician and by ChatGPT-4. Clinician and ChatGPT-4 summarisation took place November 2023-January 2024. AI and clinician-authored summaries were then rated blind by three clinicians that had seen neither that record nor summary previously.

**Figure 1:**
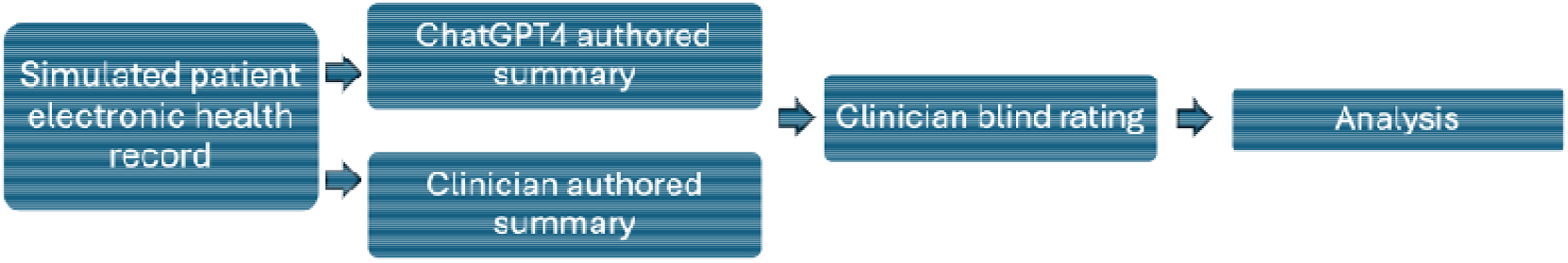
Schematic of study design.

### Expert clinician recruitment and training

A convenience sample of seven doctors working in general practice was recruited via local professional networks, all of whom are co-authors in the project. All clinicians attended two 90-minute online workshops, which included an introduction to the study and methods.

During the workshops, clinicians were given trial EHRs (authored by LS and TB) to summarise and rate, and had the opportunity to ask questions and achieve consensus on the methods of creating notes and summaries. The first workshop took place before the simulated EHR generation began. The second workshop took place before summaries were rated.

### Simulated EHR preparation

To maintain patient confidentiality when using LLMs, no real patient data was used. Instead, a sample of 70 simulated patient EHRs were generated by the clinicians based on profiles designed to use as a template to ensure a representative and varied set of notes. Simulated patient profiles were prepared by LS, consisting of age, gender, chronic conditions and repeat medications. These profiles were designed to be representative of demographics according to data on patients registered at GP practices in England (16) and prevalence of health conditions in the population as documented in Quality and Outcomes Framework (QOF) data (see Appendix A for details). Each participant co-author was allocated ten simulated patient profiles to generate notes in free text form for the ten most recent GP-entered consultations, reflecting an average patient’s most recent two-year history of contact with their GP practice.

The clinicians attended a first workshop where a consensus approach to generating the EHRs was agreed. Any queries that arose during the writing process were circulated with responses to maintain consistency. All clinicians had current or recent experience of working in general practice, such that both the patient narratives and the writing style would be as reflective of genuine general practice as possible. Documentation styles vary according to clinicians (17) and having multiple different doctors contribute to note writing helped reflect the range of writing styles in practice. Age, gender, pre-existing conditions and repeat prescriptions were presented as lists at the start of the record, followed by the free text consultation entries. This was reflective of how the author team reviewed structured data in the EHR prior to reviewing free-text data in practice. An example of the start of the record and first free text entry can be seen below in Figure 2. Spelling errors were not corrected to improve the realism. More extensive examples can be seen in Appendix B.

**Figure 2:**
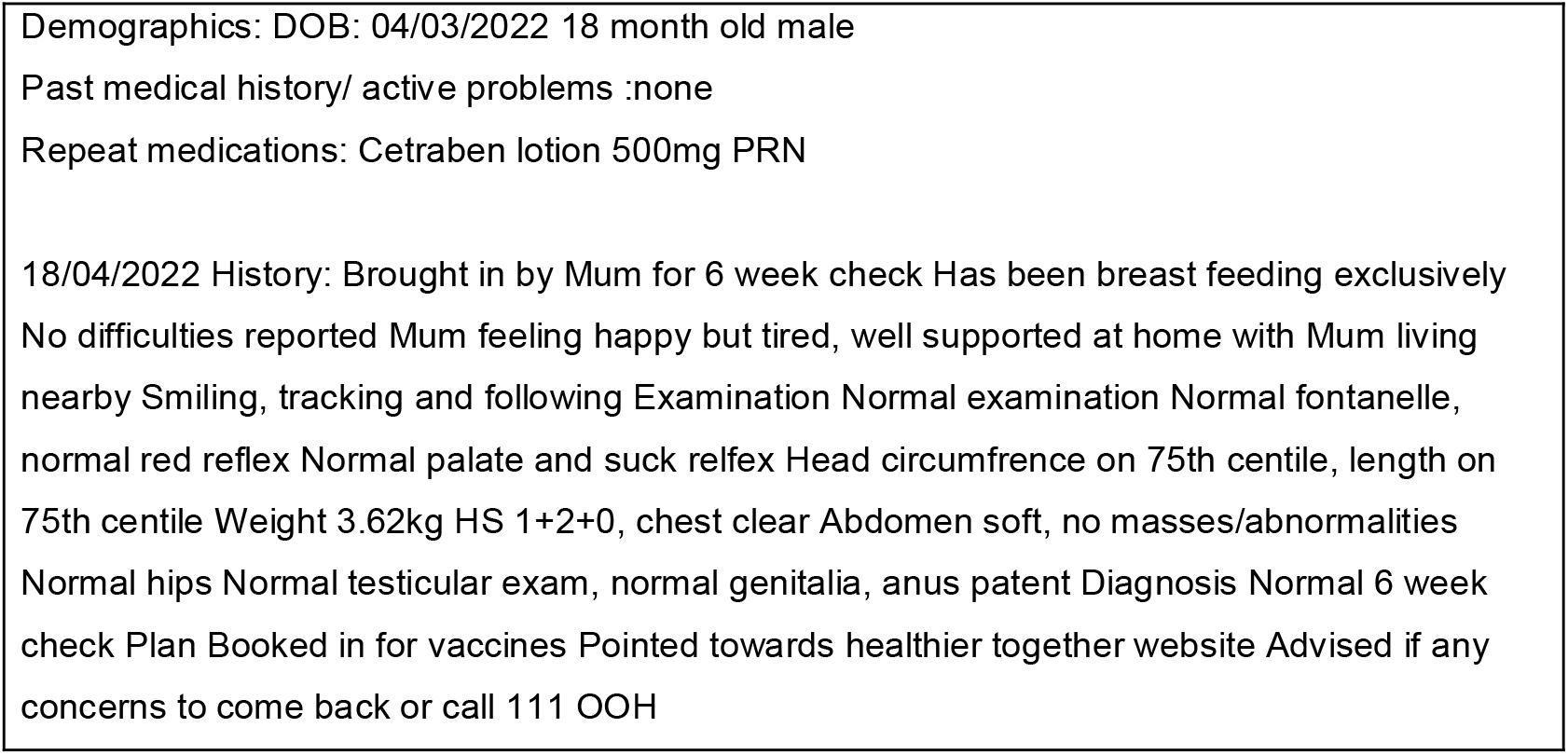
Example template and first (of ten) free text entries for a simulated patient record.

### ChatGPT-4-authored summaries

ChatGPT version 4 (ChatGPT-4, used with default settings)(18) was asked to summarise each simulated EHR using the prompt *“This is an example simulated patient record. Please summarise the important points into a paragraph of approximately 100 words intended for a clinician to read before seeing the patient”*. This prompt was followed by the simulated EHR (structured as in Figure 2). Two simulated EHRs that were also used in the study were used in a pilot. The two EHRs were selected to represent a range of complexity, one set being from a young patient with a less complex history, and another from an older patient with more comorbidities. Before the study, we piloted six different prompts, three of which evolved from the best of an initial three prompts trialed. The prompt used for the study returned more usable and consistent summaries of those tested (Appendix B).

### Clinician-authored summaries

Each clinician was assigned at random to summarise each of ten EHRs (excluding those EHRs which they generated), with an identical briefing to that used for AI: *“This is an example simulated patient record. Please summarise the important points into a paragraph of approximately 100 words intended for a clinician to read before seeing the patient”*. Clinicians were also asked to document the time taken to read through and assimilate the information in the EHR, prior to writing the summary.

### Review of summaries

Each simulated EHR and associated clinician and AI-authored summaries were assigned at random to three clinicians (excluding clinicians who had generated the notes or summary). Clinicians were also given access to the full simulated EHR to allow accuracy to be assessed. For each set of EHR notes, the two summaries (AI and parallel clinician-authored summary) were independently rated by the three clinicians, resulting in each clinician rating 60 summaries in total (30 AI-authored and 30 clinician-authored). Clinicians were not given information on whether the summary was clinician or AI-authored.

The survey questions were designed to capture multiple domains deemed relevant to a high-quality summary. Each summary was rated on a five-point Likert scale (1-strongly agree, 2-somewhat agree, 3-neither agree nor disagree, 4-somewhat disagree, 5-strongly disagree) in answer to the following eight statements. These were developed from criteria reported in an earlier study, with additional criteria based on input from the study team during the second methodology workshop (12).

1. I understand this summary
2. This summary includes relevant information
3. This summary omitted important information
4. This summary is useful to other clinicians
5. I can accept this summary without edits and language used is acceptable to share with a patient
6. This summary does not include superfluous information
7. This summary contains inaccuracies
8. This summary may contribute to bias (either regarding protected characteristics or clinical bias)

In addition, clinicians were asked to provide an overall rating of each summary from 0-10, 10 being the highest score. The question they were asked was “Overall, how would you rate this summary out of 10, 10 being the highest score?” Clinicians were also asked whether they thought the summary was generated by a human or AI. An additional question provided space for free text comments on each summary which were analysed qualitatively when interrogating reasons for differences in ratings between AI- and clinician-authored summaries for a particular simulated EHR.

### Statistical analysis

Overall ratings for each clinician- and AI-authored summary were compared and we present the minimum, mean and maximum rating for each summary for comparison. A non-parametric Wilcoxon signed rank test was used to assess whether differences in the median ratings between AI- and clinician-authored summaries were statistically significant (p<0.05). We calculated the pairwise differences for the minimum, mean and maximum overall rating for each clinician- and AI-authored summary to examine the extent of variation of overall rating across clinician-vs AI-authored summaries. In addition, we identified the clinician- and AI-authored summaries whose authorship had been misclassified by the clinicians in the ratings process.

### Ethics

As this project does not use any human participants or data, ethics approval was not required. All participant clinicians were co-authors on the project.

### Results

A total of 70 patient EHRs were simulated by seven primary care clinicians. Each EHR was summarised by one clinician and by ChatGPT-4, leading to 140 summaries. Each summary was reviewed by three different clinicians, leading to 420 responses (Figure 1). See Appendix A for a breakdown of the demographic and chronic condition profiles of the simulated patients.

### Time taken to read and assimilate information contained in simulated EHRs

The time taken to read and assimilate information contained in simulated EHRs was documented for 60 out of 70 clinician-authored summaries (one clinician did not record the time taken). The median time taken was 7 minutes 8 seconds, with a range from 1 minute 25 seconds to 20 minutes and interquartile range of 6 minutes 24 seconds.

### Overall ratings of summaries

Clinicians rated clinician-authored summaries higher than AI-authored summaries (mean score: 7.39 versus 7.00; p=0.02). There was a larger variability in how clinicians rated clinician-authored summaries (mean pairwise distance (2.04)) than AI-authored summaries (mean pairwise distance (1.38)) Figure 3 and Appendix C). Clinicians were able to correctly identify the author of the summaries as a clinician or AI 89.8% of the time (Table 1). 16 (7.6%) AI-authored summaries were incorrectly attributed to clinicians and 10 (4.8%) clinician-authored records were incorrectly attributed to AI. Clinicians were unsure of the author of clinician-authored records in 8 cases (3.8%) and of AI-authored records in 9 cases (4.3%).

**Table 1:**
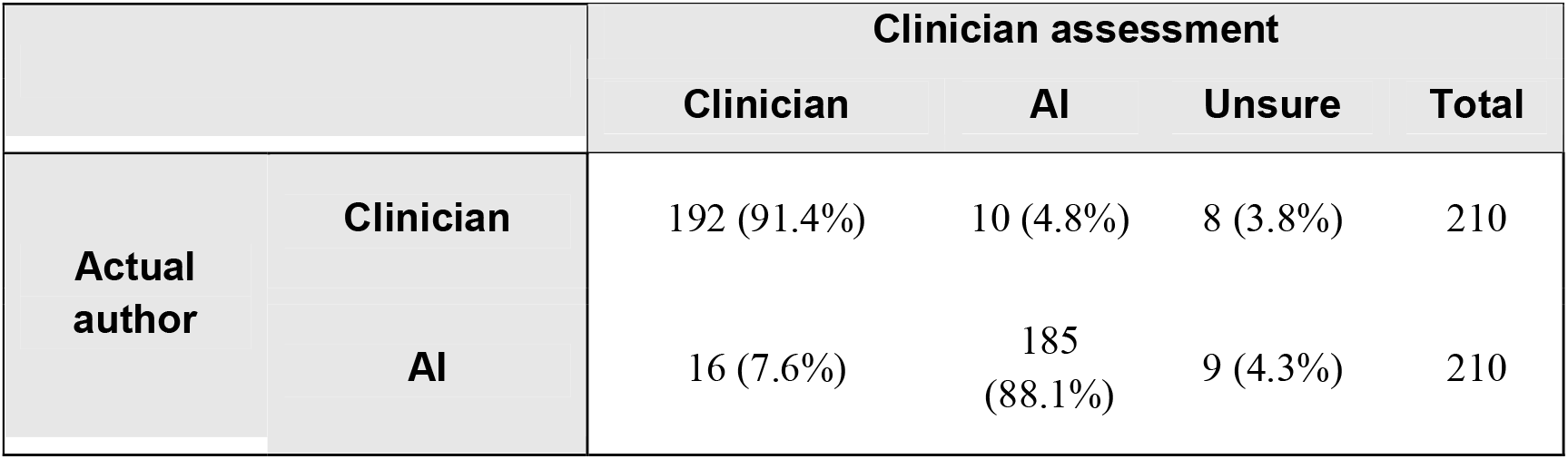
Presumed author of summary compared with actual authors.

|  |  | Clinician assessment |  |  |  |
| --- | --- | --- | --- | --- | --- |
|  |  | Clinician | AI | Unsure | Total |
| Actual author | Clinician | 192 (91.4%) | 10 (4.8%) | 8 (3.8%) | 210 |
|  | AI | 16 (7.6%) | 185 (88.1%) | 9 (4.3%) | 210 |

**Figure 3:**
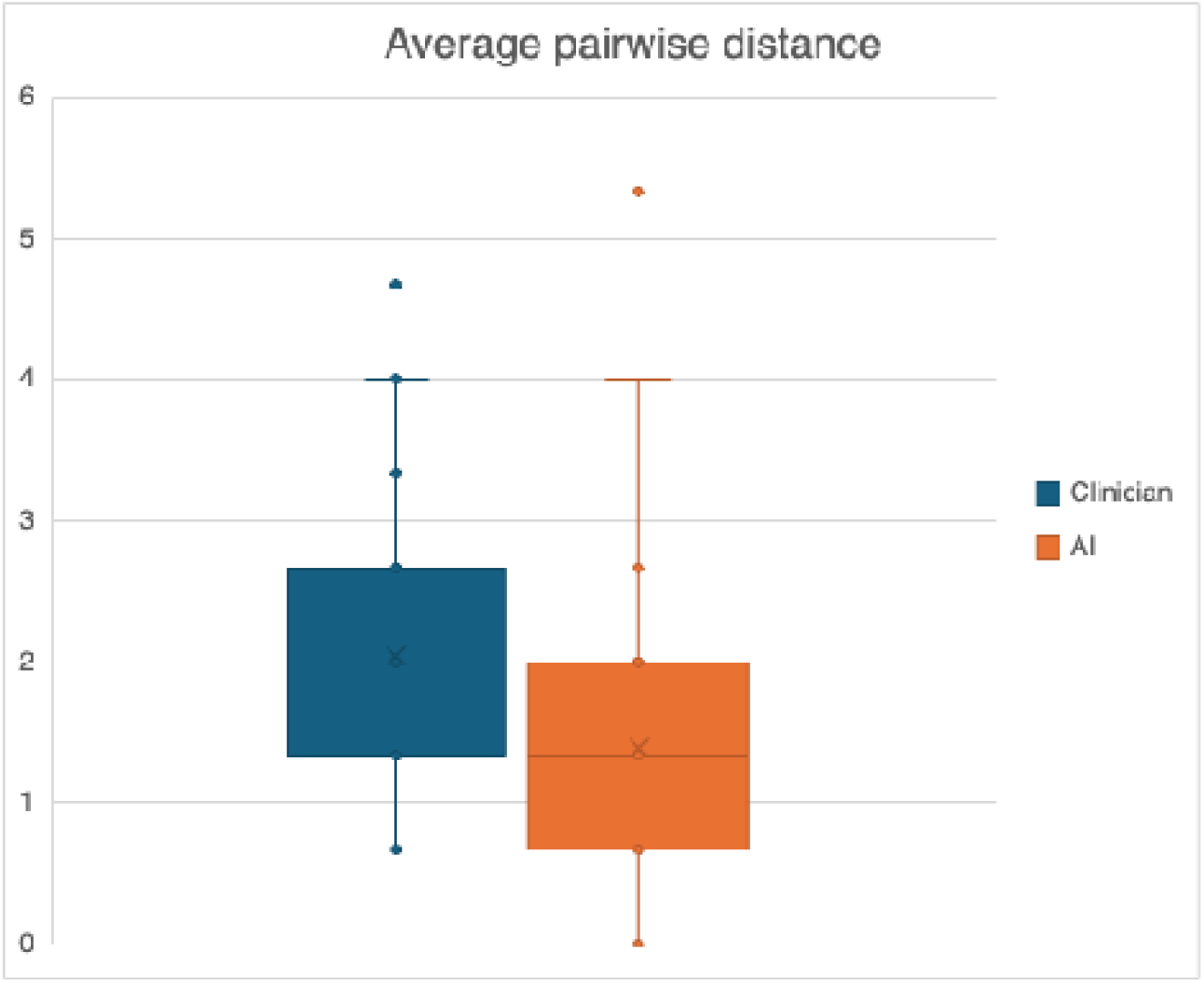
Pairwise distances between the three ratings assigned by clinicians for clinician and AI-authored summaries.

### Quality domains of summaries

Clinician-authored summaries were rated higher in including relevant information and not including superfluous information (Figure 4). In almost all cases (96.7%), clinicians strongly agreed or somewhat agreed with the statement ‘This summary includes relevant information’ for clinician-authored summaries, compared to 92.9% for AI-authored summaries (Figure 4). AI-authored summaries were perceived to contain more superfluous information. 91.9% of clinicians strongly agreed or somewhat agreed with the statement ‘This summary does not include superfluous information’ for clinician-authored summaries, compared to 40.5% for AI-authored summaries. In contrast, AI-authored summaries were rated higher than clinician-authored summaries in avoiding omission of important information (Figure 4).

**Figure 4.**
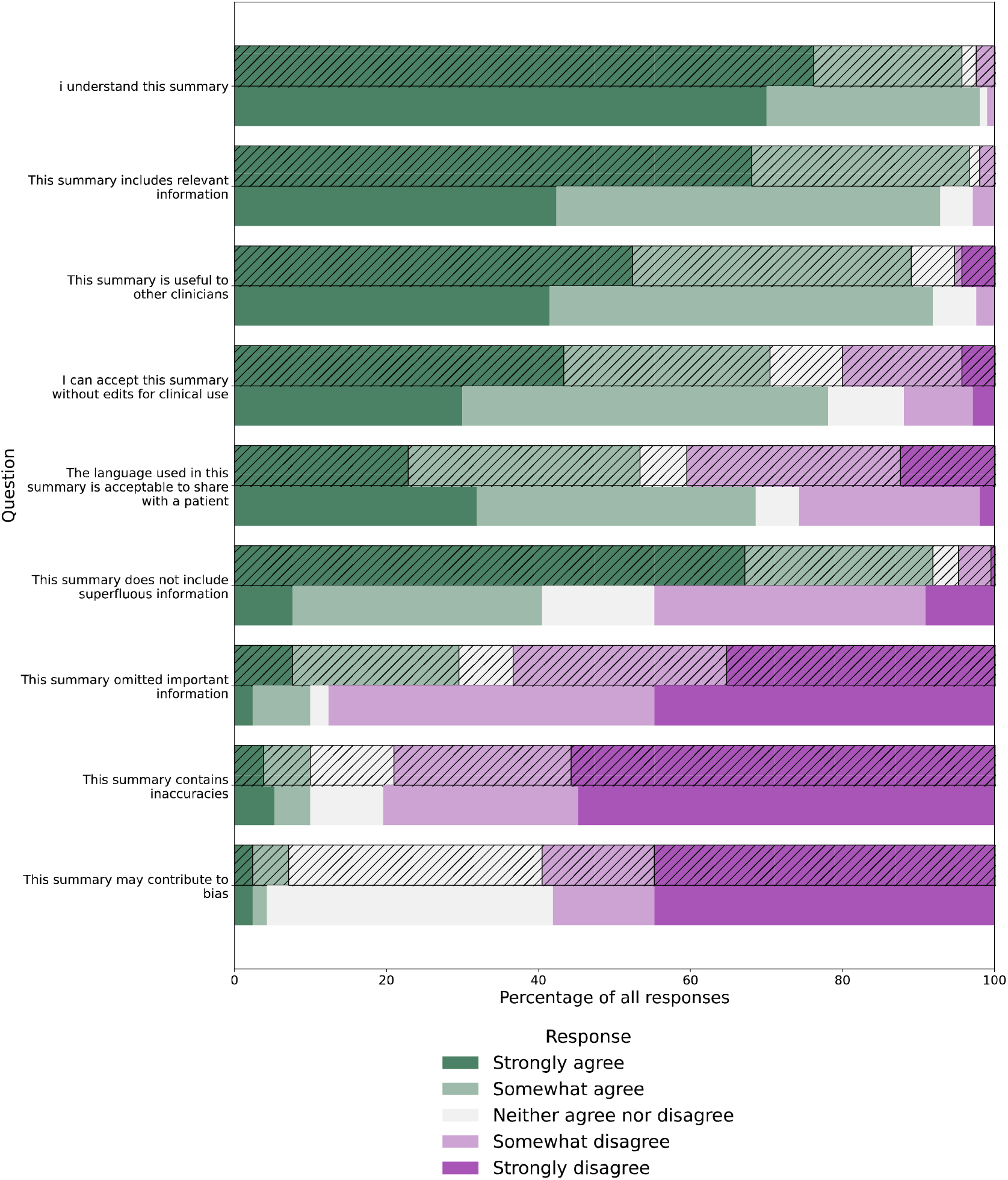
Combined figure of all responses (cross-hatched bars are clinician-authored, plain are AI-authored)

Overall, more clinicians strongly agreed or somewhat agreed with the statements ‘I understand this summary’, ‘This summary is useful to other clinicians’ and ‘I can accept this summary without edits for clinical use’ for AI-authored than clinician-authored summaries. Inaccuracies and bias were perceived to be rare in both clinician- and AI-authored summaries, with little difference between the two (Figure 4). Generally, AI-authored summaries were rated higher than clinician-authored summaries in using language acceptable to share with a patient (Figure 4). 68.6% of clinicians strongly agreed or somewhat agreed with the statement ‘The language used in this summary is acceptable to share with a patient’ for AI-authored summaries compared to 53.3% for clinician-authored summaries (Figure 4).

### Qualitative assessment of summaries

We examined cases where the mean overall rating of the AI-authored summaries was greater than that of the clinician-authored summaries (this happened in 25 out of 70 cases). This revealed consistency in the qualities that AI performed better in compared to clinicians overall, such as not omitting important information and acceptability of language to share with a patient. Generally, clinicians found the AI-authored summaries were clear, concise, chronological, neutral, relevant, accurate and readable while some clinicians found the corresponding clinician-authored summaries lacked detail and context and in one case made a significant omission.

We also interrogated examples where raters identified inaccuracies, incorrect information or hallucination in the summaries (clinician- or AI-authored). This revealed 11 discrete incidences where clinician raters detected hallucination, inaccuracies or incorrect information in AI-authored summaries. Of these, only one was labelled as a hallucination by the rater. In this instance, the AI-authored summary included a “*decision to escalate diabetes medication due to worsening control”*, which the rater identified as hallucination on the basis that “*the dose was increased because the patient tolerated 500mg MR Metformin - the repeat Hba1c had not been done yet”*. There were 12 discrete incidences where clinician raters detected inaccuracies or incorrect information in clinician-authored summaries, and no episodes labelled by raters as hallucination.

## Discussion

Primary care EHRs contain large amounts of patient information which is time-consuming for clinicians to review. This risks important information being missed, potentially compromising patient safety (5,6). Our study demonstrates the feasibility of using a generic LLM (ChatGPT-4) to create summaries of patient EHR notes to support clinicians that are overall comparable to the clinician-authored summaries across the domains of quality assessed. Summaries authored by AI were overall rated by clinicians only slightly lower than summaries authored by clinicians (7.00 out of 10, versus 7.39). Clinicians found clinician-authored summaries contained more relevant information and were less likely to include superfluous information, but AI-authored summaries were easier to understand and more acceptable for use, both by clinicians and patients, without edits. Importantly for patient safety in clinical practice, AI-authored summaries were less likely to omit important information than clinician-authored summaries. Finding AI-authored summaries were more likely to contain language acceptable to share with patients suggests their potential use in patient-facing tools for summarising medical notes. Demonstrating the potential of LLM summarisation to transform clinical workloads, the median time taken for a clinician to read through the patient record before summarising, was 7 minutes 8 seconds, which is over 70% of the standard 10-minute appointment time in UK general practice (19).

The finding that clinician- and AI-authored summaries were rated similarly in accuracy and bias provides evidence towards the potential of AI to undertake this task for clinicians in practice. This is important given the concerns with hallucination and bias that have been reported in different contexts, for example in clinical decision-making tools (20). In the context of clinical note summarisation, some research suggests hallucination may relate to lack of explicit detail in the notes (21). In our study, interrogating the qualitative data submitted by raters revealed a similar incidence of references to inaccuracies or incorrect information in AI-authored vs clinician-authored summaries, and one incident where a rater identified hallucination in an AI-authored summary. Bias is not unique to AI and decision-making by clinical staff may also be biased by factors such as protected characteristics (22-24). Ultimately, technology that outperforms humans is the goal. However, in the context of the significant time taken for clinicians to undertake this task, parity in performance may be enough to justify implementation, allowing staff to spend a higher proportion of the consultation on direct patient care.

AI-authored summaries were found to include more patient-friendly language, an important finding which suggests AI’s potential use for patient-facing summaries of information in EHRs. Future research could explore which specific features of summaries clinicians and patients value most. Understanding these preferences and reaching consensus on quality criteria will be crucial in optimising AI tools to complement clinical workflows and ensure their safe and effective integration into healthcare practice.

### Comparison with existing literature

The evidence on the competency of different AI technologies in completing different clinical tasks is rapidly evolving. For example, research demonstrates LLMs can match the level of a specialist clinician in making a diagnosis (25-27). Recent research has shown the potential of ChatGPT and other LLMs in clinical summarisation tasks, from clinical dialogues to radiology reports, with LLMs performing well and even outperforming clinicians in some contexts (13–15). Given the lack of objective or gold-standard metrics for assessing the quality of summaries, other studies have adopted the same approach as we used, with expert clinician evaluation of summaries (13).

Earlier work has found limitations in the ability of LLMs to perform specific tasks relevant to summarisation, such as accounting for the chronological ordering of events (28). A recent study found an LLM could transform discharge summaries into a format understandable to patients, though not consistently without omissions or inaccuracies (29). Heterogeneity of research in both the specific technology and clinical context limits comparability between studies (30). To our knowledge, our study is the only published research comparing clinician evaluation of clinician versus AI-authored summaries of simulated primary care EHRs. With AI being increasingly deployed in healthcare workflows, EHR note summarisation could be a vanguard use-case for time-saving in healthcare organisations such as the NHS (31).

### Strengths and limitations

A strength of this study is that seven active clinicians experienced in primary care generated simulated patient notes, designed to be as representative as possible of a real population in general practice in England. However, a limitation of the study is that despite the steps taken to make these EHRs as realistic as possible, simulated EHRs may not fully capture the complexity and variability of real patient data, including complex clinical narratives and missing information. This may affect the generalisability of AI models trained on simulated data and their performance in real-world clinical settings. Future work using real patient notes within a secure data environment would enhance the generalisability of findings to real-world health settings.

Another strength of the study is that summaries were rated by clinicians who have worked in primary care. Yet, while clinicians assessed the quality of the summaries for clinical use, the effect of using the summaries in practice is unknown and we need to better understand the clinical significance of differences between AI-generated and clinician-generated summaries. Future research could assess the value of summaries on clinical endpoints, such as studying the impact of AI-authored summaries on patient safety errors.

On average, clinicians took over 7 minutes to read and assimilate the information in the patient notes, which is unlikely to be realised in practice in the context of standard 10-minute GP appointments. Therefore, the quality of summaries authored by clinicians for the purpose of this study may be higher than what is achievable in practice. This could mean that AI-authored summaries would consistently outperform clinician-authored summaries in the context of a live general practice environment. There was more consistency between ratings for AI-authored than clinician-authored summaries which may reflect clinicians curating summaries to their own personal preferences. The finding that AI-authored summaries that perceived as more consistent by clinicians supports the potential of AI to provide a standardised summary for use in clinical practice.

In this study, clinicians were given both the AI and clinician-authored summaries for each set of EHR notes and could accurately discern whether the summary had been generated by a clinician or AI in over 90% of cases. This is high compared to detection rates reported in other settings(32,33). This may confer bias towards higher scores for clinician summaries if clinicians perceive themselves to be better than AI at summarisation and are therefore more likely to award a higher rating.

### Implications for practice

We found that a general purpose LLM, ChatGPT-4, can produce summaries of patient notes which are similarly accurate to those generated by clinicians, with potentially significant time-saving benefits for clinical practice. Such summaries could be read by clinicians to understand a patient’s relevant medical history before a consultation, or to include as a background history alongside a referral to another clinician. LLM summarisation could reduce demands on primary care clinicians and in doing so, address risk factors for work-related stress (35) that are currently contributing to challenges with workforce retention (36). An important area for further research is determining the impact of implementing LLM summarisation on clinician workload and wellbeing in practice.

However, such technology is not yet ready for implementation. Further research building on ours should aim to determine whether the results demonstrated in this study can be replicated when using real, rather than simulated, patient EHRs. This should be performed within a secure data environment complying with information governance and ethics standards. This should include an evaluation of any systematic algorithmic bias in the use of AI tools (34).

Furthermore, we used a general purpose LLM, but there is scope to improve the performance of the LLM specifically for summarisation by fine-tuning a model on high quality examples of summaries, and by refining the prompts used as input to the LLM.

## Conclusions

In this study, we demonstrated the use of a general purpose LLM (ChatGPT-4) to generate concise summaries of free text consultation histories from simulated primary care EHR data that perform similarly across multiple domains of quality compared to summaries authored by clinicians. Although AI-authored summaries were rated slightly lower than clinician-authored summaries overall, they were more consistent, less likely to omit important information and had comparable accuracy to clinician-authored summaries. Given the substantial time taken for clinicians to read and assimilate information recorded in EHRs, AI summarisation tools have the potential to save clinicians time, particularly as these tools are further enhanced, enabling clinicians to spend more time on direct patient care and to reduce errors resulting from missed information. Further evaluation is needed to better understand the features that define a high-quality summary and to assess performance using real patient notes before such technology is implemented in practice. Nevertheless, our findings suggest the feasibility, utility and acceptability of using AI-authored summaries to integrate into EHRs to support clinicians in primary care. By automating tasks, improving clinical decision-making, and enhancing communication, AI has the potential to significantly improve healthcare productivity, leading to better patient outcomes and more efficient healthcare systems.

## Data Availability

All data produced in the present study are available upon reasonable request to the authors

## Acknowledgements

At the time of this work, LS, AS, CQ, JM and MM were supported by a National Institute for Health Research (NIHR) Academic Clinical Fellowship. AN and AM were supported by the NIHR Applied Research Collaboration Northwest London. TB acknowledges support from the Wellcome Trust. The views expressed in this publication are those of the authors and not necessarily those of the NHS, the NIHR, the Department of Health and Social Care or the Wellcome Trust.

