## Appendix A for "Comparing AI- versus clinician-authored summaries of simulated primary care electronic health records"

**Appendix A- Quality and Outcomes Framework (QOF)**

The Quality and Outcomes Framework (QOF) is a voluntary annual reward and incentive programme for all GP practices in England. Practices are awarded points for achieving healthcare delivery in different domains, including in relation to certain healthcare conditions. In this study, data collected for the QOF programme has been used as an indicator of prevalence of common healthcare conditions amongst the population registered with GP practices in England, as means of creating a representative cohort of simulated patient notes.

The QOF prevalence of chronic conditions that was reflected across the 70 simulated EHRs is listed below

| Condition | Prevalence (%) |
| --- | --- |
| Asthma | 6.5 |
| Atrial fibrillation | 2.1 |
| Cancer | 3.3 |
| Chronic Kidney Disease | 4.0 |
| Chronic Obstructive Pulmonary Disease | 1.9 |
| Dementia | 0.7 |
| Depression | 12.7 |
| Diabetes Mellitus | 7.3 |
| Epilepsy | 0.7 |
| Heart failure | 1.o |
| Hypertension | 14.0 |
| Learning disability | 0.6 |
| Mental health | 0.1 |
| Non diabetic hyperglycaemia | 6.1 |
| Obesity | 9.7 |
| Osteoporosis | 0.9 |
| Palliative care | 0.5 |
| Peripheral arterial disease | 0.6 |
| Rheumatoid arthritis | 0.8 |
| Secondary prevention of coronary heart disease | 3.0 |
| Stroke and TIA | 1.8 |

The demographic profiles of the 70 simulated patients followed this distrubtion, taken from NHS data on patients registered at a GP practice in England (12)

|  | Female (%) | Male (%) |
| --- | --- | --- |
| 0-9 | 5.1 | 5.4 |
| 10-19 | 5.7 | 5.9 |
| 20-29 | 6.5 | 6.5 |
| 30-39 | 7.3 | 7.7 |
| 40-49 | 6.3 | 6.8 |
| 50-59 | 6.5 | 6.7 |
| 60-69 | 5.4 | 5.3 |
| 70-79 | 4.3 | 3.9 |
| 80-89 | 2.2 | 1.7 |
| 90+ | 0.6 | 0.3 |
