## Appendix B for "Comparing AI- versus clinician-authored summaries of simulated primary care electronic health records"

**Appendix B - Methods: Prompts**

Three preliminary prompts were trialed using a small sample of sets of simulated patient notes generated by LS. These were

●	This is an example patient primary care record. Produce a maximum 100 word summary paragraph to brief a clinician that does not know the patient’s history, covering the important aspects of the patient’s history. Less relevant information can be excluded from the summary.

●	Summarise this example patient’s record into a maximum 150 word handover prose paragraph containing the pertinent information to brief a clinician on the patient’s history.

●	This is an example simulated patient record. Please summarise the important points into a paragraph of maximum 100 words.

LS and TB screened the outputs from the prompts trialed. The prompt that was a most reflective summary of the simulated patient notes and most clinically useful was

●	This is an example simulated patient record. Please summarise the important points into a paragraph of approximately 100 words

At the initial meeting with participant co-authors, some of the co-authors queried which purpose the summary should be directed for. It was agreed that identify this purpose would be important for how both human and ChatGPT-4 generated summaries should be rated, for example a summary intended to justify a referral to a specialist service would be ‘angled’ differently to a general summary. It was agreed that for the purposes of the study, the summaries generated should be general summaries, useful for clinicians reading the patient’s notes prior to reviewing them, that could also serve as context for a referral rather than being specifically angled for this purpose. For this reason LS, TB and JC edited the prompt to specify this context to

●	This is an example simulated patient record. Please summarise the important points into a paragraph of approximately 100 words intended for a clinician to read before seeing the patient

LS, TB and JC then moved to trialing this prompt with two sets of simulated patient notes generated by the participant co authors. The two sets of notes were selected to represent a range of complexity, one set being from a young patient with a less complex history, and another from an older patient with more comorbidities.

The prompt was edited to explore whether asking ChatGPT-4 for ‘a paragraph of approximately 100 words’ was compromising the quality of the summaries.

We compared summaries generated by the following prompts on patient two

●	A) This is an example simulated patient record. Please summarise the important points into a paragraph of approximately 100 words intended for a clinician to read before seeing the patient

●	B) This is an example simulated patient record. Please summarise the important points intended for a clinician to read before seeing the patient.

●	C) This is an example simulated patient record. Please summarise the important points into a paragraph intended for a clinician to read before seeing the patient

LS, TB and JC agreed that a concise summary paragraph was more concise and clinically useful than condensed bullet points. (Summary by prompt A (A1) more concise clinically useful that prompt B (B1)).

LS, TB and JC did not find that A (asking for approximately 100 words) differed significantly in clinical utility vs C (no word limit) (A1 vs C1). However, when trialled with a more complex simulated patient record (patient 79) we found including ‘approximately 100 words’ in the prompt generated a more concise summary of similar content (A2 vs C2).

Therefore we decided to proceed with prompt A

●	A) This is an example simulated patient record. Please summarise the important points into a paragraph of approximately 100 words intended for a clinician to read before seeing the patient

We then fed ChatGPT-4 prompt A with patient 79’s simulated notes in three separate threads and though noted some differences, we found the results broadly consistent:

A2 vs A3 vs A4.

We therefore decided to proceed with prompt A for the remainder of the project.

A1

This is an example simulated patient record. Please summarise the important points into a paragraph of approximately 100 words intended for a clinician to read before seeing the patient. Demographics: DOB: 04/03/2022 18 month old male Past medical history: / active problems none Repeat medications: Cetraben lotion 500mg PRN 18/04/2022 History Brought in by Mum for 6 week check Has been breast feeding exclusively No difficulties reported Mum feeling happy but tired, well supported at home with Mum living nearby Smiling, tracking and following Examination Normal examination Normal fontanelle, normal red reflex Normal palate and suck relfex Head circumfrence on 75th centile, length on 75th centile Weight 3.62kg HS 1+2+0, chest clear Abdomen soft, no masses/abnormalities Normal hips Normal testicular exam, normal genitalia, anus patent Diagnosis Normal 6 week check Plan Booked in for vaccines Pointed towards healthier together website Advised if any concerns to come back or call 111 OOH 03/05/22 History Redness on leg following first set of vaccines yesterday Small area of redness surrounding site No discharge, does not appear painful, not spreading No temperatures Normal feeding, normal stools, normal wet nappy Examination Legs reviewed, small area of erythema at injection site, less than one cm Quite dry Does not appear, tender, no signs of infection Diagnosis Sensitive skin, no red flags identified Plan Reassurance given Advised to monitor – if redness spreading/any discharge/baby appears more irritable/less intake/decreased wet nappies/parental concern to call back or 111 OOH 11/05/22 History Brought in by Mum to review nappy rash Developed recently since changing nappy size to larger size so changing less frequently Baby well Examination Child well Dry skin across body Nappy rash across buttocks Diagnosis Dry skin Nappy rash Plan Advice re prevention Cetraben daily for dry skin 18/05/22 History Mum concerned nappy rash still not improving Baby remains well Dry skin improving overall Examination Nappy rash still present but improved Diagnosis Resolving nappy rash Plan Explained re barrier creams otc Continue emollients 19/09/22 History Mum concerned re change in stool colour and amount Normally stools seedy, loose, yellow, 4 times a day Had one episode of not going for one week, then returned to normal Recently tried first solid foods Still breast feeding alongside Has managed sucking on bread, seems to like taste Tried cooked carrot yesterday Since then stool occasionally darker in colour, more yellow-green, harder, more formed Baby well, babbling, well in himself, playing normally No signs of pain or distress Examination Looks clinically well, happy smiley baby Normal observations, normal weight HS 1+2+0, chest clear Abdomen soft, non tender Diagnosis Stool change likely due to introduction of solids Plan Reassurance given Advised stools will change with introduction of new foods Explained signs of constipation/distress to look out for 20/10/22 History Skin flares when eating hazelnuts Develops urticarial rash Photos reviewed Examination Well baby No rash currently Diagnosis ? Hazelnut intolerance Plan Avoid hazelnuts currently Introduce other nuts carefully Paediatric allergy referral 01/11/22 History Coryzal, fevers over past 1 day Temperature up to 38, goes down with calpol Feeding normally, normal nappies Examination Looks coryzal Normal observations Chest clear Diagnosis Viral URTI Plan Monitor TCB if worsening/not improving 03/11/22 History Ongoing fevers up and down Developed increased WOB yesterday Decreased intake since then, less than 50% normal intake Worsening Examination Increased RR, normal sats Nasal flaring, intercostal recssion Diagnosis Bronchiolitis Plan Referred to Paediatric A+E 5/12/2022 History Went to A+E early November Has a cold again Mum worried Due to go traveling until summer 2023 Examination Normal obs, normal exam Happy well baby Diagnosis Viral URTI, no red flags Plan Advised to get vaccines at 1 year Reassurance given re URTI 03/07/23 History Recently back from travelling Needs more cetraban Skin remains good Wasn’t able to return for one year vaccines Wants to be rebooked Due to start Nursery in September when Mum goes back to work Examination - Diagnosis Plan Booked in for delayed one year vaccines Cetraben issued – advised can request on repeat

ChatGPT-4

An 18-month-old male has a clear past medical history. At his 6-week check, he was found to be healthy with breastfeeding going well. He experienced minor skin issues post-vaccination and a nappy rash that took some time to resolve. He has shown signs of a potential hazelnut intolerance, developing an urticarial rash after consumption. The child had an episode of bronchiolitis in November 2022, which required a referral to Paediatric A+E. The child recently returned from traveling and has missed his one-year vaccines. He requires Cetraben lotion for skin care and will be starting nursery soon. The mother is seeking to rebook his missed vaccines.

B1

This is an example simulated patient record. Please summarise the important points intended for a clinician to read before seeing the patient. Demographics: DOB: 04/03/2022 18 month old male Past medical history: / active problems none Repeat medications: Cetraben lotion 500mg PRN 18/04/2022 History Brought in by Mum for 6 week check Has been breast feeding exclusively No difficulties reported Mum feeling happy but tired, well supported at home with Mum living nearby Smiling, tracking and following Examination Normal examination Normal fontanelle, normal red reflex Normal palate and suck relfex Head circumfrence on 75th centile, length on 75th centile Weight 3.62kg HS 1+2+0, chest clear Abdomen soft, no masses/abnormalities Normal hips Normal testicular exam, normal genitalia, anus patent Diagnosis Normal 6 week check Plan Booked in for vaccines Pointed towards healthier together website Advised if any concerns to come back or call 111 OOH 03/05/22 History Redness on leg following first set of vaccines yesterday Small area of redness surrounding site No discharge, does not appear painful, not spreading No temperatures Normal feeding, normal stools, normal wet nappy Examination Legs reviewed, small area of erythema at injection site, less than one cm Quite dry Does not appear, tender, no signs of infection Diagnosis Sensitive skin, no red flags identified Plan Reassurance given Advised to monitor – if redness spreading/any discharge/baby appears more irritable/less intake/decreased wet nappies/parental concern to call back or 111 OOH 11/05/22 History Brought in by Mum to review nappy rash Developed recently since changing nappy size to larger size so changing less frequently Baby well Examination Child well Dry skin across body Nappy rash across buttocks Diagnosis Dry skin Nappy rash Plan Advice re prevention Cetraben daily for dry skin 18/05/22 History Mum concerned nappy rash still not improving Baby remains well Dry skin improving overall Examination Nappy rash still present but improved Diagnosis Resolving nappy rash Plan Explained re barrier creams otc Continue emollients 19/09/22 History Mum concerned re change in stool colour and amount Normally stools seedy, loose, yellow, 4 times a day Had one episode of not going for one week, then returned to normal Recently tried first solid foods Still breast feeding alongside Has managed sucking on bread, seems to like taste Tried cooked carrot yesterday Since then stool occasionally darker in colour, more yellow-green, harder, more formed Baby well, babbling, well in himself, playing normally No signs of pain or distress Examination Looks clinically well, happy smiley baby Normal observations, normal weight HS 1+2+0, chest clear Abdomen soft, non tender Diagnosis Stool change likely due to introduction of solids Plan Reassurance given Advised stools will change with introduction of new foods Explained signs of constipation/distress to look out for 20/10/22 History Skin flares when eating hazelnuts Develops urticarial rash Photos reviewed Examination Well baby No rash currently Diagnosis ? Hazelnut intolerance Plan Avoid hazelnuts currently Introduce other nuts carefully Paediatric allergy referral 01/11/22 History Coryzal, fevers over past 1 day Temperature up to 38, goes down with calpol Feeding normally, normal nappies Examination Looks coryzal Normal observations Chest clear Diagnosis Viral URTI Plan Monitor TCB if worsening/not improving 03/11/22 History Ongoing fevers up and down Developed increased WOB yesterday Decreased intake since then, less than 50% normal intake Worsening Examination Increased RR, normal sats Nasal flaring, intercostal recssion Diagnosis Bronchiolitis Plan Referred to Paediatric A+E 5/12/2022 History Went to A+E early November Has a cold again Mum worried Due to go traveling until summer 2023 Examination Normal obs, normal exam Happy well baby Diagnosis Viral URTI, no red flags Plan Advised to get vaccines at 1 year Reassurance given re URTI 03/07/23 History Recently back from travelling Needs more cetraban Skin remains good Wasn’t able to return for one year vaccines Wants to be rebooked Due to start Nursery in September when Mum goes back to work Examination - Diagnosis Plan Booked in for delayed one year vaccines Cetraben issued – advised can request on repeat

ChatGPT-4

**Summary for Clinician:**

**Demographics:**
18 month old male, DOB: 04/03/2022

**Past Medical History:**

●	No active problems.

**Medications:**

●	Cetraben lotion 500mg PRN.

**Visit Highlights:**

1. **18/04/2022:** Normal 6-week check. Baby is breastfeeding exclusively, growing well (75th centile for head circumference and length). Booked in for vaccines.
2. **03/05/2022:** Small redness on leg after vaccines, no signs of infection. Sensitive skin diagnosis. Advised to monitor.
3. **11/05/2022:** Nappy rash observed. Prescribed Cetraben daily.
4. **18/05/2022:** Nappy rash improving, but still present. Advised to use barrier creams OTC and continue emollients.
5. **19/09/2022:** Stool change after introduction of solid foods (bread and carrot). Reassured this is normal with introduction of solids.
6. **20/10/2022:** Urticarial rash after eating hazelnuts. Possible hazelnut intolerance diagnosed. Paediatric allergy referral made.
7. **01/11/2022:** Coryzal with fevers. Diagnosed with a viral URTI.
8. **03/11/2022:** Increased respiratory rate, nasal flaring, decreased feeding. Diagnosed with bronchiolitis and referred to Paediatric A+E.
9. **5/12/2022:** Had a cold post A+E visit. Travel plans until summer 2023. Reassured about URTI and advised on 1-year vaccines.
10. **03/07/23:** Recently returned from travel. Requested more Cetraben. Delayed one-year vaccines needed. Starting nursery in September.

**Recommendations:**

●	Book for delayed one-year vaccines.

●	Issue Cetraben and add to repeat prescription.

●	Monitor any allergic reactions, especially related to nuts.

●	Reassure the mother and provide guidance on introducing new foods and managing common baby health concerns.

C1

This is an example simulated patient record. Please summarise the important points into a paragraph intended for a clinician to read before seeing the patient. Demographics: DOB: 04/03/2022 18 month old male Past medical history: / active problems none Repeat medications: Cetraben lotion 500mg PRN 18/04/2022 History Brought in by Mum for 6 week check Has been breast feeding exclusively No difficulties reported Mum feeling happy but tired, well supported at home with Mum living nearby Smiling, tracking and following Examination Normal examination Normal fontanelle, normal red reflex Normal palate and suck relfex Head circumfrence on 75th centile, length on 75th centile Weight 3.62kg HS 1+2+0, chest clear Abdomen soft, no masses/abnormalities Normal hips Normal testicular exam, normal genitalia, anus patent Diagnosis Normal 6 week check Plan Booked in for vaccines Pointed towards healthier together website Advised if any concerns to come back or call 111 OOH 03/05/22 History Redness on leg following first set of vaccines yesterday Small area of redness surrounding site No discharge, does not appear painful, not spreading No temperatures Normal feeding, normal stools, normal wet nappy Examination Legs reviewed, small area of erythema at injection site, less than one cm Quite dry Does not appear, tender, no signs of infection Diagnosis Sensitive skin, no red flags identified Plan Reassurance given Advised to monitor – if redness spreading/any discharge/baby appears more irritable/less intake/decreased wet nappies/parental concern to call back or 111 OOH 11/05/22 History Brought in by Mum to review nappy rash Developed recently since changing nappy size to larger size so changing less frequently Baby well Examination Child well Dry skin across body Nappy rash across buttocks Diagnosis Dry skin Nappy rash Plan Advice re prevention Cetraben daily for dry skin 18/05/22 History Mum concerned nappy rash still not improving Baby remains well Dry skin improving overall Examination Nappy rash still present but improved Diagnosis Resolving nappy rash Plan Explained re barrier creams otc Continue emollients 19/09/22 History Mum concerned re change in stool colour and amount Normally stools seedy, loose, yellow, 4 times a day Had one episode of not going for one week, then returned to normal Recently tried first solid foods Still breast feeding alongside Has managed sucking on bread, seems to like taste Tried cooked carrot yesterday Since then stool occasionally darker in colour, more yellow-green, harder, more formed Baby well, babbling, well in himself, playing normally No signs of pain or distress Examination Looks clinically well, happy smiley baby Normal observations, normal weight HS 1+2+0, chest clear Abdomen soft, non tender Diagnosis Stool change likely due to introduction of solids Plan Reassurance given Advised stools will change with introduction of new foods Explained signs of constipation/distress to look out for 20/10/22 History Skin flares when eating hazelnuts Develops urticarial rash Photos reviewed Examination Well baby No rash currently Diagnosis ? Hazelnut intolerance Plan Avoid hazelnuts currently Introduce other nuts carefully Paediatric allergy referral 01/11/22 History Coryzal, fevers over past 1 day Temperature up to 38, goes down with calpol Feeding normally, normal nappies Examination Looks coryzal Normal observations Chest clear Diagnosis Viral URTI Plan Monitor TCB if worsening/not improving 03/11/22 History Ongoing fevers up and down Developed increased WOB yesterday Decreased intake since then, less than 50% normal intake Worsening Examination Increased RR, normal sats Nasal flaring, intercostal recssion Diagnosis Bronchiolitis Plan Referred to Paediatric A+E 5/12/2022 History Went to A+E early November Has a cold again Mum worried Due to go traveling until summer 2023 Examination Normal obs, normal exam Happy well baby Diagnosis Viral URTI, no red flags Plan Advised to get vaccines at 1 year Reassurance given re URTI 03/07/23 History Recently back from travelling Needs more cetraban Skin remains good Wasn’t able to return for one year vaccines Wants to be rebooked Due to start Nursery in September when Mum goes back to work Examination - Diagnosis Plan Booked in for delayed one year vaccines Cetraben issued – advised can request on repeat

ChatGPT-4

The patient is an 18-month-old male with no significant past medical history. He's been using Cetraben lotion PRN for dry skin issues. At his 6-week check, he was exclusively breastfeeding and showed normal growth and developmental milestones. Subsequent visits included concerns about redness post-vaccination, nappy rash, and stool changes after introducing solid foods. Notably, the patient showed a potential hazelnut intolerance characterized by an urticarial rash. Later, he experienced a viral URTI followed by bronchiolitis, which required a referral to Paediatric A+E. After traveling, the patient returned recently and was booked in for delayed one-year vaccines. The mother also requested more Cetraben for his skin. The child is set to start Nursery in September.

A2

This is an example simulated patient record. Please summarise the important points into a paragraph of approximately 100 words intended for a clinician to read before seeing the patient. Demographics: DOB: 18/05/1962 Past medical history: / active problems Hypertension Secondary prevention of coronary heart disease Heart Failure MI Repeat medications: Amlodipine 5mg OD Ramipril 2.5mg OD Ezetimibe 10mg OD Clopidogrel 75mg OD Furosemide 20mg OD 1 (10/02/22) History Telephone call on duty list Woke up at 8am this morning with central crushing chest pain Time is now 8:30, constant, radiating to jaw, severity 8/10 associated SOB, nausea and dizziness known HTN and smoker father died of MI age 65 Examination - Diagnosis ? ACS, within PCI window Plan Advised need to attend A&E to rule out heart attack Called 999 on patients behalf Called back to confirm patient received by ambulance crew 25 mins later 2 (18/02/22) History TC with patient on duty list Hospital discharge summary reviewed from Docman Initial Troponin 380 ECG: anterior STEMI stent to LAD Discharged from hospital 5 days later with secondary prevention medications Feeling okay since discharge Quite shocked over diagnosis Wanted explanation of new medications given to him Discussed rationale for each medication prescribed Has follow up with cardiology in 6 weeks Discussed lifestyle measurement including weight loss, healthy eating and exercise Examination - Diagnosis MI – anterior STEMI, stent to LAD Plan To call 999 if develops any cardiac symptoms including chest pain, SOB, palpitations Has follow up with cardiology in 6 weeks 3 (20/02/22) History TC with patient on duty list Started on Atorvastatin 80mg 2 days ago as part of secondary prevention Gradually developed muscle pain especially in arms and legs No other side effects noted No jaundice Examination - Diagnosis Reaction to statins Plan Stop statins for now Face to face exam Check LFTs, U+Es, CK Consider other statin or other lipid lower medication once bloods back 4 (25/02/22) History Face to face exam Muscle pain shortly after starting 80mg Atorvastatin CK of 700 from 50, ALT 110 from 30, other bloods including LFTs normal Examination BP: 139/92 A – own, patent B – chest clear, no crackles, no wheeze, sats 99% room air, RR 18 C – HS: I+I+0, pulse regular and strong, CRT 2 seconds, calves SNT, HR 70, temp 37.0 D – GCS 15/15, Pupils: PEARL E – abdomen SNT, BS present No jaundice Diagnosis Statin intolerance – explained that statins are affecting liver function and will need to stop Plan Continue to withhold statins for now Continue other regular medication for now A&G Cardiology Dear Cardiology, this patient had an anterior STEMI with a stent to LAD on 10/02/22 and is due to be seen in your clinic in 4 weeks. He was started on Atorvastatin 80mg and developed myalgia 3 days later. Bloods revealed a CK of 700 from 50, ALT 110 from 40, other bloods including LFTs normal. I have advised him to stop taking Atorvastatin for now. Would you recommend another statin therapy or alternative lipid lower medication? 5 (01/03/22) History TC with patient See previous consultation re statin intolerance A&G from Cardiology: if no contraindications, please switch to Ezetimibe 10mg OD and we will see him in clinic shortly Discussed Cardiology recommendations – patient happy with plan to switch to Ezetimibe 10mg OD Went through potential side effects of medication Examination - Diagnosis - Plan Stop Atorvastatin 80mg OD Start Ezetimibe 10mg OD 6 (04/03/22) History Patient unable to request new medication Medications only in acute section Recent hospital admission for STEMI Examination - Diagnosis - Plan Issue below medication – add to regular medication Amlodipine 5mg OD Ramipril 2.5mg OD Ezetimibe 10mg OD Clopidogrel 75mg OD 7 (14/04/23) History Telephone call with patient had an anterior STEMI with a stent to LAD on 10/02/23 3-month history of increasing SOB when walking up stairs 1-month history of bilateral ankle swelling Generally more fatigued than usual No chest pain, palpitations, PND No cough, wheeze, sputum, hemoptysis No fever, rigors, weight loss Compliant with medication Under cardiology, next review in 6 months Examination ? Heart failure 2nd MI Diagnosis Plan Face to face exam Echo ECG Bloods including BNP today Referral to HF nurse if BNP raised 8 (15/04/23) History Face to face review See previous consultation for history BNP 1500, all other bloods normal ECG - NSR Examination Bilateral pitting oedema in ankles A – own, patent B – chest clear, no crackles, no wheeze, sats 96% room air, RR 16 C – HS: I+I+0, pulse regular and strong, CRT 2 seconds, calves SNT, BP 141/90, HR 60, temp 36.8 D – GCS 15/15 E – abdomen SNT, BS present Diagnosis Likely heart failure with evidence of fluid overload (pitting oedema in ankles) Plan Expedite echo Start 20 furosemide OD Repeat U+Es in 1 week and face to face review To call Dr if develops worsening fluid overload (ankle swelling, SOB, cough, chest pain, palpitations, dizziness) 9 (25/04/23) History See previous consultation Repeat U+Es normal, no evidence of AKI Passing urine normally – good urine output Echo: heart failure with preserved ejection fraction, LVEF 56% Explained echo results to patient and need to take water tablets Examination Ankles improved – minimal pitting oedema A – own, patent B – chest clear, no crackles, no wheeze, sats 97% room air, RR 18 C – HS: I+I+0, pulse regular and strong, CRT 2 seconds, calves SNT, BP 139/94, HR 65, temp 36.5 D – GCS 15/15 E – abdomen SNT, BS present Diagnosis heart failure with preserved ejection fraction, LVEF 56% fluid overload improving Plan Continue 20 Furosemide – tolerating well To call Dr if develops worsening fluid overload (ankle swelling, SOB, cough, chest pain, palpitations, dizziness) Referral to HF nurse with BNP and echo results Dear HF nurse, this patient had an anterior STEMI with a stent to LAD on 10/02/22 and is under Cardiology. He developed HF symptoms 3 months ago, BNP was 1,500 and echo showed HFpEF with LVEF 56%. I have started him on 20mg Furosemide and he has responded well to this. Please could you advise on next steps in terms of medication and follow up? 10 (20/05/23) History MI with HFpEF and LVED 56% Reply from heart failure nurse team: please continue on 20mg Furosemide and the heart failure team will see him in clinic in 6 weeks time HF nurse advice explained to patient Continues to tolerate 20 Furosemide well No signs of AKI or further fluid overload Ankles now have minimal swelling Examination - Diagnosis - Plan Continue 20 Furosemide – tolerating well To call Dr if develops worsening fluid overload (ankle swelling, SOB, cough, chest pain, palpitations, dizziness) HF team review in 6 weeks time

ChatGPT-4

The patient, born on 18/05/1962, has a medical history significant for hypertension, coronary heart disease, heart failure, and myocardial infarction (MI). In February 2022, the patient presented with symptoms suggestive of acute coronary syndrome, confirmed as anterior STEMI, leading to a stent placement in the LAD. Following the MI, he was prescribed secondary prevention medications including Atorvastatin, which was later discontinued due to muscle pain and elevated blood markers. The patient was switched to Ezetimibe based on cardiology recommendations. By April 2023, he reported increasing shortness of breath and bilateral ankle swelling, raising suspicions of heart failure. Investigations revealed heart failure with preserved ejection fraction (HFpEF) with an LVEF of 56%. Treatment with Furosemide was initiated with improvements noted. The heart failure team plans to review him in 6 weeks.

C2

This is an example simulated patient record. Please summarise the important points into a paragraph intended for a clinician to read before seeing the patient. Demographics: DOB: 18/05/1962 Past medical history: / active problems Hypertension Secondary prevention of coronary heart disease Heart Failure MI Repeat medications: Amlodipine 5mg OD Ramipril 2.5mg OD Ezetimibe 10mg OD Clopidogrel 75mg OD Furosemide 20mg OD 1 (10/02/22) History Telephone call on duty list Woke up at 8am this morning with central crushing chest pain Time is now 8:30, constant, radiating to jaw, severity 8/10 associated SOB, nausea and dizziness known HTN and smoker father died of MI age 65 Examination - Diagnosis ? ACS, within PCI window Plan Advised need to attend A&E to rule out heart attack Called 999 on patients behalf Called back to confirm patient received by ambulance crew 25 mins later 2 (18/02/22) History TC with patient on duty list Hospital discharge summary reviewed from Docman Initial Troponin 380 ECG: anterior STEMI stent to LAD Discharged from hospital 5 days later with secondary prevention medications Feeling okay since discharge Quite shocked over diagnosis Wanted explanation of new medications given to him Discussed rationale for each medication prescribed Has follow up with cardiology in 6 weeks Discussed lifestyle measurement including weight loss, healthy eating and exercise Examination - Diagnosis MI – anterior STEMI, stent to LAD Plan To call 999 if develops any cardiac symptoms including chest pain, SOB, palpitations Has follow up with cardiology in 6 weeks 3 (20/02/22) History TC with patient on duty list Started on Atorvastatin 80mg 2 days ago as part of secondary prevention Gradually developed muscle pain especially in arms and legs No other side effects noted No jaundice Examination - Diagnosis Reaction to statins Plan Stop statins for now Face to face exam Check LFTs, U+Es, CK Consider other statin or other lipid lower medication once bloods back 4 (25/02/22) History Face to face exam Muscle pain shortly after starting 80mg Atorvastatin CK of 700 from 50, ALT 110 from 30, other bloods including LFTs normal Examination BP: 139/92 A – own, patent B – chest clear, no crackles, no wheeze, sats 99% room air, RR 18 C – HS: I+I+0, pulse regular and strong, CRT 2 seconds, calves SNT, HR 70, temp 37.0 D – GCS 15/15, Pupils: PEARL E – abdomen SNT, BS present No jaundice Diagnosis Statin intolerance – explained that statins are affecting liver function and will need to stop Plan Continue to withhold statins for now Continue other regular medication for now A&G Cardiology Dear Cardiology, this patient had an anterior STEMI with a stent to LAD on 10/02/22 and is due to be seen in your clinic in 4 weeks. He was started on Atorvastatin 80mg and developed myalgia 3 days later. Bloods revealed a CK of 700 from 50, ALT 110 from 40, other bloods including LFTs normal. I have advised him to stop taking Atorvastatin for now. Would you recommend another statin therapy or alternative lipid lower medication? 5 (01/03/22) History TC with patient See previous consultation re statin intolerance A&G from Cardiology: if no contraindications, please switch to Ezetimibe 10mg OD and we will see him in clinic shortly Discussed Cardiology recommendations – patient happy with plan to switch to Ezetimibe 10mg OD Went through potential side effects of medication Examination - Diagnosis - Plan Stop Atorvastatin 80mg OD Start Ezetimibe 10mg OD 6 (04/03/22) History Patient unable to request new medication Medications only in acute section Recent hospital admission for STEMI Examination - Diagnosis - Plan Issue below medication – add to regular medication Amlodipine 5mg OD Ramipril 2.5mg OD Ezetimibe 10mg OD Clopidogrel 75mg OD 7 (14/04/23) History Telephone call with patient had an anterior STEMI with a stent to LAD on 10/02/23 3-month history of increasing SOB when walking up stairs 1-month history of bilateral ankle swelling Generally more fatigued than usual No chest pain, palpitations, PND No cough, wheeze, sputum, hemoptysis No fever, rigors, weight loss Compliant with medication Under cardiology, next review in 6 months Examination ? Heart failure 2nd MI Diagnosis Plan Face to face exam Echo ECG Bloods including BNP today Referral to HF nurse if BNP raised 8 (15/04/23) History Face to face review See previous consultation for history BNP 1500, all other bloods normal ECG - NSR Examination Bilateral pitting oedema in ankles A – own, patent B – chest clear, no crackles, no wheeze, sats 96% room air, RR 16 C – HS: I+I+0, pulse regular and strong, CRT 2 seconds, calves SNT, BP 141/90, HR 60, temp 36.8 D – GCS 15/15 E – abdomen SNT, BS present Diagnosis Likely heart failure with evidence of fluid overload (pitting oedema in ankles) Plan Expedite echo Start 20 furosemide OD Repeat U+Es in 1 week and face to face review To call Dr if develops worsening fluid overload (ankle swelling, SOB, cough, chest pain, palpitations, dizziness) 9 (25/04/23) History See previous consultation Repeat U+Es normal, no evidence of AKI Passing urine normally – good urine output Echo: heart failure with preserved ejection fraction, LVEF 56% Explained echo results to patient and need to take water tablets Examination Ankles improved – minimal pitting oedema A – own, patent B – chest clear, no crackles, no wheeze, sats 97% room air, RR 18 C – HS: I+I+0, pulse regular and strong, CRT 2 seconds, calves SNT, BP 139/94, HR 65, temp 36.5 D – GCS 15/15 E – abdomen SNT, BS present Diagnosis heart failure with preserved ejection fraction, LVEF 56% fluid overload improving Plan Continue 20 Furosemide – tolerating well To call Dr if develops worsening fluid overload (ankle swelling, SOB, cough, chest pain, palpitations, dizziness) Referral to HF nurse with BNP and echo results Dear HF nurse, this patient had an anterior STEMI with a stent to LAD on 10/02/22 and is under Cardiology. He developed HF symptoms 3 months ago, BNP was 1,500 and echo showed HFpEF with LVEF 56%. I have started him on 20mg Furosemide and he has responded well to this. Please could you advise on next steps in terms of medication and follow up? 10 (20/05/23) History MI with HFpEF and LVED 56% Reply from heart failure nurse team: please continue on 20mg Furosemide and the heart failure team will see him in clinic in 6 weeks time HF nurse advice explained to patient Continues to tolerate 20 Furosemide well No signs of AKI or further fluid overload Ankles now have minimal swelling Examination - Diagnosis - Plan Continue 20 Furosemide – tolerating well To call Dr if develops worsening fluid overload (ankle swelling, SOB, cough, chest pain, palpitations, dizziness) HF team review in 6 weeks time

ChatGPT-4

The patient, born on 18/05/1962, has a past medical history of hypertension, coronary heart disease secondary prevention, heart failure, and myocardial infarction (MI). On 10/02/22, the patient presented with chest pain radiating to the jaw, shortness of breath, nausea, and dizziness and was suspected to have acute coronary syndrome (ACS). Emergency services were alerted and the patient was confirmed to have an anterior STEMI, leading to a stent being placed in the LAD. Subsequent hospitalization revealed an initial troponin level of 380 and the patient was discharged on secondary prevention medications. However, upon initiation of Atorvastatin 80mg, the patient developed muscle pain and elevated CK and ALT levels, indicating a reaction to the statin. Atorvastatin was ceased and, following cardiology advice, the patient was switched to Ezetimibe 10mg OD. On 14/04/23, the patient reported increasing shortness of breath, ankle swelling, and fatigue over several months. Elevated BNP levels and an echocardiogram showing heart failure with preserved ejection fraction (LVEF 56%) confirmed a diagnosis of heart failure likely secondary to the MI. Treatment with Furosemide 20mg was started and showed improvement in symptoms. A heart failure nurse team recommended continuation of Furosemide and scheduled a review in 6 weeks.

A3

This is an example simulated patient record. Please summarise the important points into a paragraph of approximately 100 words intended for a clinician to read before seeing the patient. Demographics: DOB: 18/05/1962 Past medical history: / active problems Hypertension Secondary prevention of coronary heart disease Heart Failure MI Repeat medications: Amlodipine 5mg OD Ramipril 2.5mg OD Ezetimibe 10mg OD Clopidogrel 75mg OD Furosemide 20mg OD 1 (10/02/22) History Telephone call on duty list Woke up at 8am this morning with central crushing chest pain Time is now 8:30, constant, radiating to jaw, severity 8/10 associated SOB, nausea and dizziness known HTN and smoker father died of MI age 65 Examination - Diagnosis ? ACS, within PCI window Plan Advised need to attend A&E to rule out heart attack Called 999 on patients behalf Called back to confirm patient received by ambulance crew 25 mins later 2 (18/02/22) History TC with patient on duty list Hospital discharge summary reviewed from Docman Initial Troponin 380 ECG: anterior STEMI stent to LAD Discharged from hospital 5 days later with secondary prevention medications Feeling okay since discharge Quite shocked over diagnosis Wanted explanation of new medications given to him Discussed rationale for each medication prescribed Has follow up with cardiology in 6 weeks Discussed lifestyle measurement including weight loss, healthy eating and exercise Examination - Diagnosis MI – anterior STEMI, stent to LAD Plan To call 999 if develops any cardiac symptoms including chest pain, SOB, palpitations Has follow up with cardiology in 6 weeks 3 (20/02/22) History TC with patient on duty list Started on Atorvastatin 80mg 2 days ago as part of secondary prevention Gradually developed muscle pain especially in arms and legs No other side effects noted No jaundice Examination - Diagnosis Reaction to statins Plan Stop statins for now Face to face exam Check LFTs, U+Es, CK Consider other statin or other lipid lower medication once bloods back 4 (25/02/22) History Face to face exam Muscle pain shortly after starting 80mg Atorvastatin CK of 700 from 50, ALT 110 from 30, other bloods including LFTs normal Examination BP: 139/92 A – own, patent B – chest clear, no crackles, no wheeze, sats 99% room air, RR 18 C – HS: I+I+0, pulse regular and strong, CRT 2 seconds, calves SNT, HR 70, temp 37.0 D – GCS 15/15, Pupils: PEARL E – abdomen SNT, BS present No jaundice Diagnosis Statin intolerance – explained that statins are affecting liver function and will need to stop Plan Continue to withhold statins for now Continue other regular medication for now A&G Cardiology Dear Cardiology, this patient had an anterior STEMI with a stent to LAD on 10/02/22 and is due to be seen in your clinic in 4 weeks. He was started on Atorvastatin 80mg and developed myalgia 3 days later. Bloods revealed a CK of 700 from 50, ALT 110 from 40, other bloods including LFTs normal. I have advised him to stop taking Atorvastatin for now. Would you recommend another statin therapy or alternative lipid lower medication? 5 (01/03/22) History TC with patient See previous consultation re statin intolerance A&G from Cardiology: if no contraindications, please switch to Ezetimibe 10mg OD and we will see him in clinic shortly Discussed Cardiology recommendations – patient happy with plan to switch to Ezetimibe 10mg OD Went through potential side effects of medication Examination - Diagnosis - Plan Stop Atorvastatin 80mg OD Start Ezetimibe 10mg OD 6 (04/03/22) History Patient unable to request new medication Medications only in acute section Recent hospital admission for STEMI Examination - Diagnosis - Plan Issue below medication – add to regular medication Amlodipine 5mg OD Ramipril 2.5mg OD Ezetimibe 10mg OD Clopidogrel 75mg OD 7 (14/04/23) History Telephone call with patient had an anterior STEMI with a stent to LAD on 10/02/23 3-month history of increasing SOB when walking up stairs 1-month history of bilateral ankle swelling Generally more fatigued than usual No chest pain, palpitations, PND No cough, wheeze, sputum, hemoptysis No fever, rigors, weight loss Compliant with medication Under cardiology, next review in 6 months Examination ? Heart failure 2nd MI Diagnosis Plan Face to face exam Echo ECG Bloods including BNP today Referral to HF nurse if BNP raised 8 (15/04/23) History Face to face review See previous consultation for history BNP 1500, all other bloods normal ECG - NSR Examination Bilateral pitting oedema in ankles A – own, patent B – chest clear, no crackles, no wheeze, sats 96% room air, RR 16 C – HS: I+I+0, pulse regular and strong, CRT 2 seconds, calves SNT, BP 141/90, HR 60, temp 36.8 D – GCS 15/15 E – abdomen SNT, BS present Diagnosis Likely heart failure with evidence of fluid overload (pitting oedema in ankles) Plan Expedite echo Start 20 furosemide OD Repeat U+Es in 1 week and face to face review To call Dr if develops worsening fluid overload (ankle swelling, SOB, cough, chest pain, palpitations, dizziness) 9 (25/04/23) History See previous consultation Repeat U+Es normal, no evidence of AKI Passing urine normally – good urine output Echo: heart failure with preserved ejection fraction, LVEF 56% Explained echo results to patient and need to take water tablets Examination Ankles improved – minimal pitting oedema A – own, patent B – chest clear, no crackles, no wheeze, sats 97% room air, RR 18 C – HS: I+I+0, pulse regular and strong, CRT 2 seconds, calves SNT, BP 139/94, HR 65, temp 36.5 D – GCS 15/15 E – abdomen SNT, BS present Diagnosis heart failure with preserved ejection fraction, LVEF 56% fluid overload improving Plan Continue 20 Furosemide – tolerating well To call Dr if develops worsening fluid overload (ankle swelling, SOB, cough, chest pain, palpitations, dizziness) Referral to HF nurse with BNP and echo results Dear HF nurse, this patient had an anterior STEMI with a stent to LAD on 10/02/22 and is under Cardiology. He developed HF symptoms 3 months ago, BNP was 1,500 and echo showed HFpEF with LVEF 56%. I have started him on 20mg Furosemide and he has responded well to this. Please could you advise on next steps in terms of medication and follow up? 10 (20/05/23) History MI with HFpEF and LVED 56% Reply from heart failure nurse team: please continue on 20mg Furosemide and the heart failure team will see him in clinic in 6 weeks time HF nurse advice explained to patient Continues to tolerate 20 Furosemide well No signs of AKI or further fluid overload Ankles now have minimal swelling Examination - Diagnosis - Plan Continue 20 Furosemide – tolerating well To call Dr if develops worsening fluid overload (ankle swelling, SOB, cough, chest pain, palpitations, dizziness) HF team review in 6 weeks time

ChatGPT-4

The patient, born on 18/05/1962, has a medical history of hypertension, coronary heart disease, heart failure, and myocardial infarction (MI). He experienced an anterior STEMI in February and had a stent placed in the LAD. After initiating Atorvastatin 80mg as secondary prevention, he developed myalgia and raised CK and ALT levels, leading to its discontinuation and the introduction of Ezetimibe 10mg. By April, he reported symptoms suggestive of heart failure, including shortness of breath and ankle swelling. Investigations revealed a BNP of 1500 and an echo demonstrated heart failure with preserved ejection fraction (HFpEF) with an LVEF of 56%. Furosemide 20mg was commenced with improvement in symptoms. A recent communication from the heart failure nurse team recommends continuation of Furosemide with a follow-up in their clinic in six weeks.

A4

This is an example simulated patient record. Please summarise the important points into a paragraph of approximately 100 words intended for a clinician to read before seeing the patient. Demographics: DOB: 18/05/1962 Past medical history: / active problems Hypertension Secondary prevention of coronary heart disease Heart Failure MI Repeat medications: Amlodipine 5mg OD Ramipril 2.5mg OD Ezetimibe 10mg OD Clopidogrel 75mg OD Furosemide 20mg OD 1 (10/02/22) History Telephone call on duty list Woke up at 8am this morning with central crushing chest pain Time is now 8:30, constant, radiating to jaw, severity 8/10 associated SOB, nausea and dizziness known HTN and smoker father died of MI age 65 Examination - Diagnosis ? ACS, within PCI window Plan Advised need to attend A&E to rule out heart attack Called 999 on patients behalf Called back to confirm patient received by ambulance crew 25 mins later 2 (18/02/22) History TC with patient on duty list Hospital discharge summary reviewed from Docman Initial Troponin 380 ECG: anterior STEMI stent to LAD Discharged from hospital 5 days later with secondary prevention medications Feeling okay since discharge Quite shocked over diagnosis Wanted explanation of new medications given to him Discussed rationale for each medication prescribed Has follow up with cardiology in 6 weeks Discussed lifestyle measurement including weight loss, healthy eating and exercise Examination - Diagnosis MI – anterior STEMI, stent to LAD Plan To call 999 if develops any cardiac symptoms including chest pain, SOB, palpitations Has follow up with cardiology in 6 weeks 3 (20/02/22) History TC with patient on duty list Started on Atorvastatin 80mg 2 days ago as part of secondary prevention Gradually developed muscle pain especially in arms and legs No other side effects noted No jaundice Examination - Diagnosis Reaction to statins Plan Stop statins for now Face to face exam Check LFTs, U+Es, CK Consider other statin or other lipid lower medication once bloods back 4 (25/02/22) History Face to face exam Muscle pain shortly after starting 80mg Atorvastatin CK of 700 from 50, ALT 110 from 30, other bloods including LFTs normal Examination BP: 139/92 A – own, patent B – chest clear, no crackles, no wheeze, sats 99% room air, RR 18 C – HS: I+I+0, pulse regular and strong, CRT 2 seconds, calves SNT, HR 70, temp 37.0 D – GCS 15/15, Pupils: PEARL E – abdomen SNT, BS present No jaundice Diagnosis Statin intolerance – explained that statins are affecting liver function and will need to stop Plan Continue to withhold statins for now Continue other regular medication for now A&G Cardiology Dear Cardiology, this patient had an anterior STEMI with a stent to LAD on 10/02/22 and is due to be seen in your clinic in 4 weeks. He was started on Atorvastatin 80mg and developed myalgia 3 days later. Bloods revealed a CK of 700 from 50, ALT 110 from 40, other bloods including LFTs normal. I have advised him to stop taking Atorvastatin for now. Would you recommend another statin therapy or alternative lipid lower medication? 5 (01/03/22) History TC with patient See previous consultation re statin intolerance A&G from Cardiology: if no contraindications, please switch to Ezetimibe 10mg OD and we will see him in clinic shortly Discussed Cardiology recommendations – patient happy with plan to switch to Ezetimibe 10mg OD Went through potential side effects of medication Examination - Diagnosis - Plan Stop Atorvastatin 80mg OD Start Ezetimibe 10mg OD 6 (04/03/22) History Patient unable to request new medication Medications only in acute section Recent hospital admission for STEMI Examination - Diagnosis - Plan Issue below medication – add to regular medication Amlodipine 5mg OD Ramipril 2.5mg OD Ezetimibe 10mg OD Clopidogrel 75mg OD 7 (14/04/23) History Telephone call with patient had an anterior STEMI with a stent to LAD on 10/02/23 3-month history of increasing SOB when walking up stairs 1-month history of bilateral ankle swelling Generally more fatigued than usual No chest pain, palpitations, PND No cough, wheeze, sputum, hemoptysis No fever, rigors, weight loss Compliant with medication Under cardiology, next review in 6 months Examination ? Heart failure 2nd MI Diagnosis Plan Face to face exam Echo ECG Bloods including BNP today Referral to HF nurse if BNP raised 8 (15/04/23) History Face to face review See previous consultation for history BNP 1500, all other bloods normal ECG - NSR Examination Bilateral pitting oedema in ankles A – own, patent B – chest clear, no crackles, no wheeze, sats 96% room air, RR 16 C – HS: I+I+0, pulse regular and strong, CRT 2 seconds, calves SNT, BP 141/90, HR 60, temp 36.8 D – GCS 15/15 E – abdomen SNT, BS present Diagnosis Likely heart failure with evidence of fluid overload (pitting oedema in ankles) Plan Expedite echo Start 20 furosemide OD Repeat U+Es in 1 week and face to face review To call Dr if develops worsening fluid overload (ankle swelling, SOB, cough, chest pain, palpitations, dizziness) 9 (25/04/23) History See previous consultation Repeat U+Es normal, no evidence of AKI Passing urine normally – good urine output Echo: heart failure with preserved ejection fraction, LVEF 56% Explained echo results to patient and need to take water tablets Examination Ankles improved – minimal pitting oedema A – own, patent B – chest clear, no crackles, no wheeze, sats 97% room air, RR 18 C – HS: I+I+0, pulse regular and strong, CRT 2 seconds, calves SNT, BP 139/94, HR 65, temp 36.5 D – GCS 15/15 E – abdomen SNT, BS present Diagnosis heart failure with preserved ejection fraction, LVEF 56% fluid overload improving Plan Continue 20 Furosemide – tolerating well To call Dr if develops worsening fluid overload (ankle swelling, SOB, cough, chest pain, palpitations, dizziness) Referral to HF nurse with BNP and echo results Dear HF nurse, this patient had an anterior STEMI with a stent to LAD on 10/02/22 and is under Cardiology. He developed HF symptoms 3 months ago, BNP was 1,500 and echo showed HFpEF with LVEF 56%. I have started him on 20mg Furosemide and he has responded well to this. Please could you advise on next steps in terms of medication and follow up? 10 (20/05/23) History MI with HFpEF and LVED 56% Reply from heart failure nurse team: please continue on 20mg Furosemide and the heart failure team will see him in clinic in 6 weeks time HF nurse advice explained to patient Continues to tolerate 20 Furosemide well No signs of AKI or further fluid overload Ankles now have minimal swelling Examination - Diagnosis - Plan Continue 20 Furosemide – tolerating well To call Dr if develops worsening fluid overload (ankle swelling, SOB, cough, chest pain, palpitations, dizziness) HF team review in 6 weeks time

ChatGPT-4

2 / 2

The patient, born on 18/05/1962, presented with a history of hypertension, coronary heart disease, heart failure, and myocardial infarction (MI). On 10/02/22, he experienced severe chest pain with associated shortness of breath, nausea, and dizziness, and was diagnosed with potential acute coronary syndrome. The patient was later confirmed to have had an anterior STEMI with a stent placed in the LAD artery. Post-hospitalization, he developed myalgia after starting Atorvastatin 80mg, resulting in its cessation and switch to Ezetimibe 10mg OD. By April 2023, he reported shortness of breath, ankle swelling, and fatigue, leading to a diagnosis of heart failure. An echocardiogram revealed heart failure with preserved ejection fraction (HFpEF) with a left ventricular ejection fraction (LVEF) of 56%. The patient was started on Furosemide 20mg OD to manage fluid overload and will have a heart failure team review in 6 weeks.

Is this conversation helpful so far?
