## Appendix C for "Comparing AI- versus clinician-authored summaries of simulated primary care electronic health records"

Graphical representation of maximum, minimum and mean overall rating for clinician vs AI summaries

Minimum maximum and mean rating for each clinician authored summary

|  | rating one | rating two | rating three | min rating | max rating | mean rating |
| --- | --- | --- | --- | --- | --- | --- |
| summary 2 | 6 | 8 | 9 | 6 | 9 | 7.66667 |
| summary 4 | 8 | 8 | 9 | 8 | 9 | 8.33333 |
| summary 5 | 8 | 7 | 7 | 7 | 8 | 7.33333 |
| summary 6 | 9 | 7 | 10 | 7 | 10 | 8.66667 |
| summary 7 | 10 | 8 | 6 | 6 | 10 | 8 |
| summary 8 | 8 | 10 | 5 | 5 | 10 | 7.66667 |
| summary 9 | 9 | 9 | 7 | 7 | 9 | 8.33333 |
| summary 11 | 9 | 8 | 10 | 8 | 10 | 9 |
| summary 14 | 9 | 7 | 9 | 7 | 9 | 8.33333 |
| summary 16 | 8 | 6 | 10 | 6 | 10 | 8 |
| summary 17 | 9 | 4 | 6 | 4 | 9 | 6.33333 |
| summary 18 | 6 | 10 | 7 | 6 | 10 | 7.66667 |
| summary 19 | 8 | 8 | 10 | 8 | 10 | 8.66667 |
| summary 20 | 10 | 10 | 4 | 4 | 10 | 8 |
| summary 21 | 2 | 4 | 6 | 2 | 6 | 4 |
| summary 22 | 9 | 9 | 5 | 5 | 9 | 7.66667 |
| summary 23 | 6 | 7 | 6 | 6 | 7 | 6.33333 |
| summary 26 | 8 | 7 | 9 | 7 | 9 | 8 |
| summary 28 | 8 | 4 | 7 | 4 | 8 | 6.33333 |
| summary 29 | 5 | 5 | 7 | 5 | 7 | 5.66667 |
| summary 30 | 6 | 9 | 9 | 6 | 9 | 8 |
| summary 31 | 5 | 10 | 7 | 5 | 10 | 7.33333 |
| summary 32 | 7 | 6 | 9 | 6 | 9 | 7.33333 |
| summary 33 | 8 | 10 | 4 | 4 | 10 | 7.33333 |
| summary 35 | 9 | 5 | 7 | 5 | 9 | 7 |
| summary 38 | 6 | 8 | 9 | 6 | 9 | 7.66667 |
| summary 40 | 4 | 5 | 9 | 4 | 9 | 6 |
| summary 41 | 7 | 5 | 9 | 5 | 9 | 7 |
| summary 42 | 10 | 6 | 9 | 6 | 10 | 8.33333 |
| summary 43 | 5 | 6 | 6 | 5 | 6 | 5.66667 |
| summary 44 | 9 | 7 | 6 | 6 | 9 | 7.33333 |
| summary 45 | 5 | 9 | 8 | 5 | 9 | 7.33333 |
| summary 46 | 7 | 7 | 8 | 7 | 8 | 7.33333 |
| summary 47 | 10 | 9 | 10 | 9 | 10 | 9.66667 |
| summary 50 | 8 | 4 | 6 | 4 | 8 | 6 |
| summary 52 | 10 | 5 | 9 | 5 | 10 | 8 |
| summary 53 | 2 | 4 | 9 | 2 | 9 | 5 |
| summary 54 | 5 | 8 | 9 | 5 | 9 | 7.33333 |
| summary 55 | 7 | 8 | 9 | 7 | 9 | 8 |
| summary 56 | 8 | 5 | 9 | 5 | 9 | 7.33333 |
| summary 57 | 6 | 4 | 7 | 4 | 7 | 5.66667 |
| summary 58 | 9 | 9 | 7 | 7 | 9 | 8.33333 |
| summary 59 | 8 | 6 | 7 | 6 | 8 | 7 |
| summary 62 | 6 | 3 | 9 | 3 | 9 | 6 |
| summary 64 | 9 | 5 | 7 | 5 | 9 | 7 |
| summary 65 | 9 | 6 | 9 | 6 | 9 | 8 |
| summary 66 | 9 | 9 | 10 | 9 | 10 | 9.33333 |
| summary 67 | 9 | 8 | 7 | 7 | 9 | 8 |
| summary 68 | 8 | 6 | 9 | 6 | 9 | 7.66667 |
| summary 69 | 8 | 6 | 7 | 6 | 8 | 7 |
| summary 70 | 8 | 7 | 8 | 7 | 8 | 7.66667 |
| summary 71 | 7 | 4 | 7 | 4 | 7 | 6 |
| summary 74 | 10 | 8 | 5 | 5 | 10 | 7.66667 |
| summary 76 | 9 | 6 | 8 | 6 | 9 | 7.66667 |
| summary 77 | 7 | 5 | 8 | 5 | 8 | 6.66667 |
| summary 78 | 10 | 8 | 8 | 8 | 10 | 8.66667 |
| summary 79 | 7 | 5 | 7 | 5 | 7 | 6.33333 |
| summary 80 | 9 | 8 | 6 | 6 | 9 | 7.66667 |
| summary 81 | 9 | 5 | 7 | 5 | 9 | 7 |
| summary 82 | 10 | 8 | 9 | 8 | 10 | 9 |
| summary 86 | 8 | 8 | 6 | 6 | 8 | 7.33333 |
| summary 88 | 8 | 8 | 5 | 5 | 8 | 7 |
| summary 89 | 7 | 9 | 7 | 7 | 9 | 7.66667 |
| summary 90 | 10 | 9 | 9 | 9 | 10 | 9.33333 |
| summary 91 | 9 | 8 | 8 | 8 | 9 | 8.33333 |
| summary 92 | 4 | 7 | 7 | 4 | 7 | 6 |
| summary 93 | 7 | 6 | 6 | 6 | 7 | 6.33333 |
| summary 94 | 8 | 8 | 7 | 7 | 8 | 7.66667 |
| summary 98 | 8 | 9 | 3 | 3 | 9 | 6.66667 |
| summary 100 | 8 | 9 | 5 | 5 | 9 | 7.33333 |

Minimum maximum and mean rating for each AI authored summary

|  | rating one | rating two | rating three | min rating | max rating | mean rating |
| --- | --- | --- | --- | --- | --- | --- |
| summary 2 | 7 | 8 | 7 | 7 | 8 | 7.33333 |
| summary 4 | 7 | 7 | 9 | 7 | 9 | 7.66667 |
| summary 5 | 7 | 8 | 8 | 7 | 8 | 7.66667 |
| summary 6 | 8 | 9 | 6 | 6 | 9 | 7.66667 |
| summary 7 | 8 | 7 | 5 | 5 | 8 | 6.66667 |
| summary 8 | 9 | 5 | 7 | 5 | 9 | 7 |
| summary 9 | 7 | 4 | 6 | 4 | 7 | 5.66667 |
| summary 11 | 8 | 7 | 7 | 7 | 8 | 7.33333 |
| summary 14 | 8 | 6 | 7 | 6 | 8 | 7 |
| summary 16 | 9 | 8 | 5 | 5 | 9 | 7.33333 |
| summary 17 | 8 | 8 | 8 | 8 | 8 | 8 |
| summary 18 | 7 | 6 | 8 | 6 | 8 | 7 |
| summary 19 | 9 | 7 | 6 | 6 | 9 | 7.33333 |
| summary 20 | 8 | 6 | 7 | 6 | 8 | 7 |
| summary 21 | 8 | 9 | 7 | 7 | 9 | 8 |
| summary 22 | 9 | 8 | 5 | 5 | 9 | 7.33333 |
| summary 23 | 7 | 7 | 7 | 7 | 7 | 7 |
| summary 26 | 5 | 7 | 6 | 5 | 7 | 6 |
| summary 28 | 7 | 8 | 6 | 6 | 8 | 7 |
| summary 29 | 9 | 8 | 9 | 8 | 9 | 8.66667 |
| summary 30 | 9 | 7 | 9 | 7 | 9 | 8.33333 |
| summary 31 | 7 | 9 | 9 | 7 | 9 | 8.33333 |
| summary 32 | 5 | 7 | 5 | 5 | 7 | 5.66667 |
| summary 33 | 9 | 9 | 7 | 7 | 9 | 8.33333 |
| summary 35 | 9 | 7 | 7 | 7 | 9 | 7.66667 |
| summary 38 | 7 | 8 | 9 | 7 | 9 | 8 |
| summary 40 | 9 | 5 | 6 | 5 | 9 | 6.66667 |
| summary 41 | 5 | 7 | 8 | 5 | 8 | 6.66667 |
| summary 42 | 8 | 8 | 7 | 7 | 8 | 7.66667 |
| summary 43 | 8 | 7 | 6 | 6 | 8 | 7 |
| summary 44 | 6 | 6 | 7 | 6 | 7 | 6.33333 |
| summary 45 | 7 | 5 | 6 | 5 | 7 | 6 |
| summary 46 | 6 | 6 | 7 | 6 | 7 | 6.33333 |
| summary 47 | 7 | 5 | 7 | 5 | 7 | 6.33333 |
| summary 50 | 9 | 8 | 8 | 8 | 9 | 8.33333 |
| summary 52 | 3 | 7 | 7 | 3 | 7 | 5.66667 |
| summary 53 | 6 | 8 | 6 | 6 | 8 | 6.66667 |
| summary 54 | 7 | 7 | 8 | 7 | 8 | 7.33333 |
| summary 55 | 5 | 8 | 9 | 5 | 9 | 7.33333 |
| summary 56 | 6 | 8 | 7 | 6 | 8 | 7 |
| summary 57 | 7 | 7 | 7 | 7 | 7 | 7 |
| summary 58 | 7 | 5 | 7 | 5 | 7 | 6.33333 |
| summary 59 | 10 | 7 | 6 | 6 | 10 | 7.66667 |
| summary 62 | 7 | 7 | 8 | 7 | 8 | 7.33333 |
| summary 64 | 7 | 8 | 7 | 7 | 8 | 7.33333 |
| summary 65 | 6 | 8 | 7 | 6 | 8 | 7 |
| summary 66 | 7 | 6 | 7 | 6 | 7 | 6.66667 |
| summary 67 | 6 | 6 | 4 | 4 | 6 | 5.33333 |
| summary 68 | 7 | 8 | 7 | 7 | 8 | 7.33333 |
| summary 69 | 7 | 7 | 5 | 5 | 7 | 6.33333 |
| summary 70 | 7 | 7 | 5 | 5 | 7 | 6.33333 |
| summary 71 | 10 | 8 | 7 | 7 | 10 | 8.33333 |
| summary 74 | 10 | 7 | 7 | 7 | 10 | 8 |
| summary 76 | 7 | 8 | 7 | 7 | 8 | 7.33333 |
| summary 77 | 7 | 7 | 8 | 7 | 8 | 7.33333 |
| summary 78 | 5 | 7 | 7 | 5 | 7 | 6.33333 |
| summary 79 | 7 | 8 | 7 | 7 | 8 | 7.33333 |
| summary 80 | 7 | 7 | 3 | 3 | 7 | 5.66667 |
| summary 81 | 6 | 6 | 7 | 6 | 7 | 6.33333 |
| summary 82 | 6 | 7 | 7 | 6 | 7 | 6.66667 |
| summary 86 | 6 | 8 | 6 | 6 | 8 | 6.66667 |
| summary 88 | 3 | 7 | 7 | 3 | 7 | 5.66667 |
| summary 89 | 7 | 6 | 6 | 6 | 7 | 6.33333 |
| summary 90 | 7 | 7 | 7 | 7 | 7 | 7 |
| summary 91 | 6 | 7 | 4 | 4 | 7 | 5.66667 |
| summary 92 | 7 | 7 | 7 | 7 | 7 | 7 |
| summary 93 | 8 | 8 | 8 | 8 | 8 | 8 |
| summary 94 | 8 | 7 | 2 | 2 | 8 | 5.66667 |
| summary 98 | 10 | 7 | 2 | 2 | 10 | 6.33333 |
| summary 100 | 6 | 8 | 7 | 6 | 8 | 7 |
